# An Elevated Birth Prevalence of Fraser Syndrome in Quebec Linked To A Founder Pathogenic Variant

**DOI:** 10.1101/2025.10.30.25336922

**Authors:** Genevieve Gagnon, Justin Pelletier, Dariel Ashton-Beaucage, Mohadese Sayahian Dekhordi, Vincent Chapdelaine, Marie-Ange Delrue, Aspasia Karalis, Daniel Taliun, John D. Rioux, Philippe M Campeau, Claude Bhérer

**Affiliations:** Department of Human Genetics, Faculty of Medicine and Health Sciences, McGill University, Montreal, Quebec, Canada; Canadian Excellence Research Chair in Genomic Medicine and Victor Phillip Dahdaleh Institute of Genomic Medicine at McGill University, Montreal, Quebec, Canada; Department of Medicine, Faculty of Medicine, Laval University, Quebec city, Quebec, Canada; Centre de recherche, Institut de Cardiologie de Montréal, Faculté de Médecine, Université de Montréal, Montréal, Québec, Canada; Department of Medicine, Faculté de médecine, Université de Montréal, Montréal, Québec, Canada; Department of Pediatrics, Division of Genetics, CHU Sainte-Justine and University of Montreal, Montreal, Quebec, Canada

**Keywords:** Fraser syndrome, French-Canadian, p.(Arg124Ter), prevalence, founder effect

## Abstract

**Purpose:** Fraser syndrome (FS) is an autosomal recessive disorder, characterized by cryptophthalmos, syndactyly, and anomalies of the respiratory and urogenital tracts. Here we estimate the birth prevalence of FS in the French-Canadian founder population of Quebec, where no prevalence has been reported to date. We also describe the phenotype of probands with FS.

**Methods:** Pathogenic allele frequency was estimated in the Quebec IBD cohort and the population-based cohort CARTaGENE (CaG), from exome sequencing (*n*=2,323), short-read genome sequencing (*n*=2,173) and genotyping array data (*n*=29,330). Phenotypic data was collected for FS probands at CHU Ste-Justine (between 2013 and 2023).

**Results:** *FRAS1* p.(Arg124Ter) was the most frequent pathogenic variant in the Quebec IBD and CaG cohorts, with frequencies 17-fold and 21-fold higher than Non-Finnish Europeans. Four French-Canadian probands were diagnosed with FS at CHU Ste-Justine, three of whom were homozygous for the variant. Birth prevalence was 0.23 per 100,000 births when predicted from the Quebec IBD cohort, 0.98 from CaG, and 1.34 from reported cases.

**Conclusion:** FS shows an elevated birth prevalence in the French-Canadian population of Quebec. We propose *FRAS1* p.(Arg124Ter) as a candidate founder pathogenic variant, informing clinical practices in this population.

## INTRODUCTION

Fraser syndrome (FS; MIM 219000) is an autosomal recessive disorder characterized by multiple congenital malformations. It is caused by pathogenic variants in three genes, *FRAS1* (HGNC:19185),^1^ *FREM2* (HGNC:25396),^2^ and *GRIP1* (HGNC:18708),^3^ which encode extracellular matrix proteins that are essential for epithelial-mesenchymal interactions.^4^ During embryogenesis, defects in epidermal adhesion and failure of the apoptosis program explain the pathogenesis of FS.^4^ Hallmark features of this syndrome include cryptophthalmos and syndactyly. In addition to these primary manifestations, FS is characterized by anomalies of the respiratory and urogenital tracts, as well as craniofacial, umbilical, anorectal, skeletal, and cardiac defects.^5^

FS can lead to both lethal and nonlethal outcomes, with expression ranging from miscarriage or stillbirth to major physical malformations.^5,6,7^ In many cases, pregnancies are terminated due to a severe prognosis following prenatal detection of major anomalies.^7^ Given the high perinatal mortality and severe malformations associated with this syndrome, early diagnosis is crucial for counselling and managing pregnancy outcomes.^7^ Because it is a congenital disorder frequently associated with fetal loss, ascertaining the true birth prevalence of FS is challenging. In Europe, the prevalence was estimated at 0.20 per 100,000 births, based on 26 patients with FS (including 7 with parental consanguinity) reported in a population of 12 million births over a 19-year period.^7^ FS prevalence likely varies worldwide due to multiple factors, such as population structure, healthcare access, and diagnostic capabilities.^8^

A better understanding of carrier rate is required to develop effective screening programs, as recommended by the American College of Medical Genetics.^9^ This is especially important in populations with a founder effect, for which identifying pathogenic variants at elevated frequencies can inform clinical practice and healthcare policies. While no prevalence has been estimated for this syndrome in Quebec, our previous work on coding variation in the French-Canadian population of Quebec revealed significant enrichment of a coding variant in the *FRAS1* gene (Bhérer et al., medRxiv 2025; preprint PMID: 40791678). Specifically, we found through an analysis of pathogenic variants in the Quebec Inflammatory Bowel Disease (IBD) cohort that the nonsense variant c.370C>T (p.Arg124Ter) in the *FRAS1* gene was present at an unusually high frequency in French Canadians (allele frequency (AF) = 1.51E-03) relative to gnomAD non-Finnish Europeans (NFE) (AF = 8.863E-05) (Bhérer et al., medRxiv 2025; preprint PMID: 40791678). Building on this finding in the current study, in all three known FS causing genes (*FRAS1*, *FREM2* and *GRIP1*) we examine the spectrum and frequency of pathogenic variants detected within the Quebec IBD cohort. To validate these observations, we then analyze data from CARTaGENE (CaG), a large population-based cohort more representative of the general population in Quebec. To assess the clinical relevance of our genetic estimates, we review cases of FS identified over the past decade at CHU Ste-Justine, a mother-child tertiary hospital covering a large proportion of Quebec’s pediatric population. We report both the genetic and clinical characteristics of affected individuals and provide birth prevalence estimates based on all three data sources. By conducting a population-based epidemiological study to complement the insights gained from a hospital-based case series, we aim to provide knowledge on the frequency and clinical presentation of this rare disorder in this population, with potential implications for clinical and genetic counselling practices in Quebec.

## MATERIALS AND METHODS

### Quebec IBD cohort

The Quebec IBD cohort includes 3,102 individuals recruited across the province of Quebec through three studies: IBDGC-Montreal, iGenoMed-MTT, and Genome Quebec-GENIZON (Bhérer et al., medRxiv 2025; preprint PMID: 40791678). Participants were patients with a diagnosed inflammatory bowel disease, as well as parental or healthy population controls, recruited primarily at university-affiliated hospitals. Here, we used summary statistics on pathogenic variants previously identified in exome data from a subset of 2,323 French-Canadian individuals unrelated to the third degree (the so-called “UNRFC subset”) (Bhérer et al., medRxiv 2025; preprint PMID: 40791678). Details about the cohort, exome sequencing and data processing were published in Bhérer et al. (medRxiv 2025; preprint PMID: 40791678).

### CARTaGENE population-based cohort

The CaG study is both a population-based biobank and a prospective cohort of 43,000 participants between the ages of 40 and 69 at recruitment.^10^ Sampling was performed proportionally to the population distribution in six metropolitan areas, which comprise more than 50% of the Quebec population. We used genotyping data available for 29,330 individuals of diverse origins, and short-read 30x genome sequencing data available for 2,173 unrelated individuals. Details on sampling, genotyping, sequencing, and variant quality control of CaG data are described in McClelland et al. (medRxiv 2025; preprint PMID: 40463523) and in the CaG website.^11^ In genotyping data, we defined a subset of European individuals unrelated to the third degree, using PRIMUS software (v1.9.0). In genome sequencing data, we used ancestry subgroups (French-Canadians, Moroccans, Haitians) defined previously mainly based on self-reported data (McClelland et al., medRxiv 2025; preprint PMID: 40463523). Variants (SNVs and InDels) were annotated using the Variant Effect Predictor (VEP) tool (version 112) with the LOFTEE plugin (version 1.0.4) to assess predicted loss-of-function (pLoF).^12,13^

### Quebec IBD and CaG ethics and participant consent

Analysis of CaG data was approved by the ethical review board from the Faculty of Medicine and Health Sciences at McGill (IRB number A07-M45-21B). Study protocols for the Quebec IBD cohort were approved by MHI Institutional Ethics Committee “Comité d’éthique de la recherche et du développement des nouvelles technologies” (IBD Genetics 2005-23 (05-813) and MTT : MP-02-2017-7170, 2017- 2202) and the Research Ethics Office of the Faculty of Medicine and Health Sciences at McGill University, Canada (IRB number A01-M04-21A).

### CHU Ste-Justine cohort

Patients diagnosed with FS at CHU Ste-Justine between 2013 and 2023 were included retrospectively. To do so, the genetics division’s genetic testing database was searched for FS diagnosis. For observed cases, the retrospective chart review was performed in accordance with the CHU Sainte-Justine IRB-approved protocol number 4072. Patient information was de-identified. FS sequencing panels were performed after the termination of pregnancy or death in all cases and identified the pathogenic variant(s). In one case (Patient 3), the family was also enrolled in a fetal malformation genomic study which also showed the variant.^14^ Post-mortem examination was performed in all cases, and patient phenotypes were reported based on clinical information provided by referring physicians. Pedigree information was collected, and no cases of consanguinity were identified among the families included in the study. The origin of parents was self-reported.

### Manual curation of variants

In the Quebec IBD cohort data, and in CaG genome sequencing data, we extracted all variants found in any transcripts of the *FRAS1, FREM2* and *GRIP1* genes. We selected variants classified as Pathogenic (P), Likely Pathogenic (LP), or with conflicting interpretations of pathogenicity according to ClinVar (accession date: August 16, 2024), and high confidence predicted loss-of-function (pLoF) variants. We performed manual curation of these variants, excluding variants with conflicting interpretations that were without P or LP submissions in ClinVar and not predicted as high confidence pLoF variants. Curated P/LP variants were retained for subsequent analyses.

### Birth prevalence estimation

Birth prevalence was obtained by dividing the number of expected or observed cases, by the number of births reported in Quebec.^15,16^ Given the high perinatal mortality associated with this syndrome, both live births and stillbirths are included in our calculations.^7^ For French-Canadian ancestry specifically, we assume that the number of births represents 80% of total births, in line with the proportion of the French speaking population in the province.^17^

To calculate the expected frequency of FS cases, we used AF from Quebec IBD cohort and the CaG genome sequencing data separately, as well as the number of births reported in 2021.^15,16^ Assuming complete penetrance, we applied Hardy-Weinberg equation *p*^2^ + 2*pq* + *q*^2^ = 1, where *q* is the sum of pathogenic AFs, and *q*^2^ the frequency of homozygotes.

Patients were identified at CHU Sainte-Justine between 2013 and 2023. To estimate birth prevalence from clinical data, we assumed the number of births to be 40% of births reported in Quebec during the 10-year period, since this proportion reflects the population coverage by this tertiary care center.^18^

### Statistical Analysis

In the Quebec IBD cohort, for each curated pathogenic variant, we used AF in the subset of unrelated French-Canadian exomes (*n* = 2,323) to compute carrier rates. In CaG genome sequencing data, we estimated AF and carrier rate of curated pathogenic variants in 2,173 unrelated individuals and in a subset of French-Canadian genetic ancestry (*n* = 1,756). For each variant, confidence intervals on AF were computed using Jeffrey’s method and all statistical analyses were performed using the RStudio statistical package (version 1.4.1106). AF in the French-Canadian subset was compared to that observed in exome sequencing data from 64,603 NFE in the gnomAD database (v.2.1.1). We applied one-sided Fisher’s Exact test to obtain the p-value and computed bias-corrected odds ratio (OR) as described in Rivas et al.^19^ Regional carrier frequencies were estimated using a subset of 24,838 unrelated individuals of European ancestry with CaG genotyping data across the six metropolitan sampling areas.

## RESULTS

### Identification of pathogenic variants

In the Quebec IBD cohort, only the *FRAS1* p.(Arg124Ter) pathogenic variant was found (allele count (AC) = 7; allele number (AN) = 4,644; AF = 1.51E-03) (*Supplementary Table S1*). Within the CaG genomes dataset, we confirmed six FS-causing variants among 43 potential PLP variants (*Supplementary Table S3*). Four were present in individuals of French-Canadian genetic ancestry (Figure 1a, c). From the number of heterozygotes, we estimated a cumulative carrier rate of 1/160 in the French-Canadian subset (AC = 11; AN = 3,512; AF = 3.13E-03), and of 1/166 overall (AC = 13; AN = 4,346; AF = 2.99E-03). Notably, eight carriers of the *FRAS1* p.(Arg124Ter) variant were identified, all French-Canadian individuals, corresponding to a carrier rate of 1/220, representing 73% (8/11) of all P/LP alleles. The three other variants in the French-Canadian subset, *FRAS1* p.(Trp449Ter), p.(Arg1244Ter), and p.(Ser3607Ter) each had a carrier rate of 1/2,173 (AC = 1, AF = 2.85E-04).

**Figure 1.**
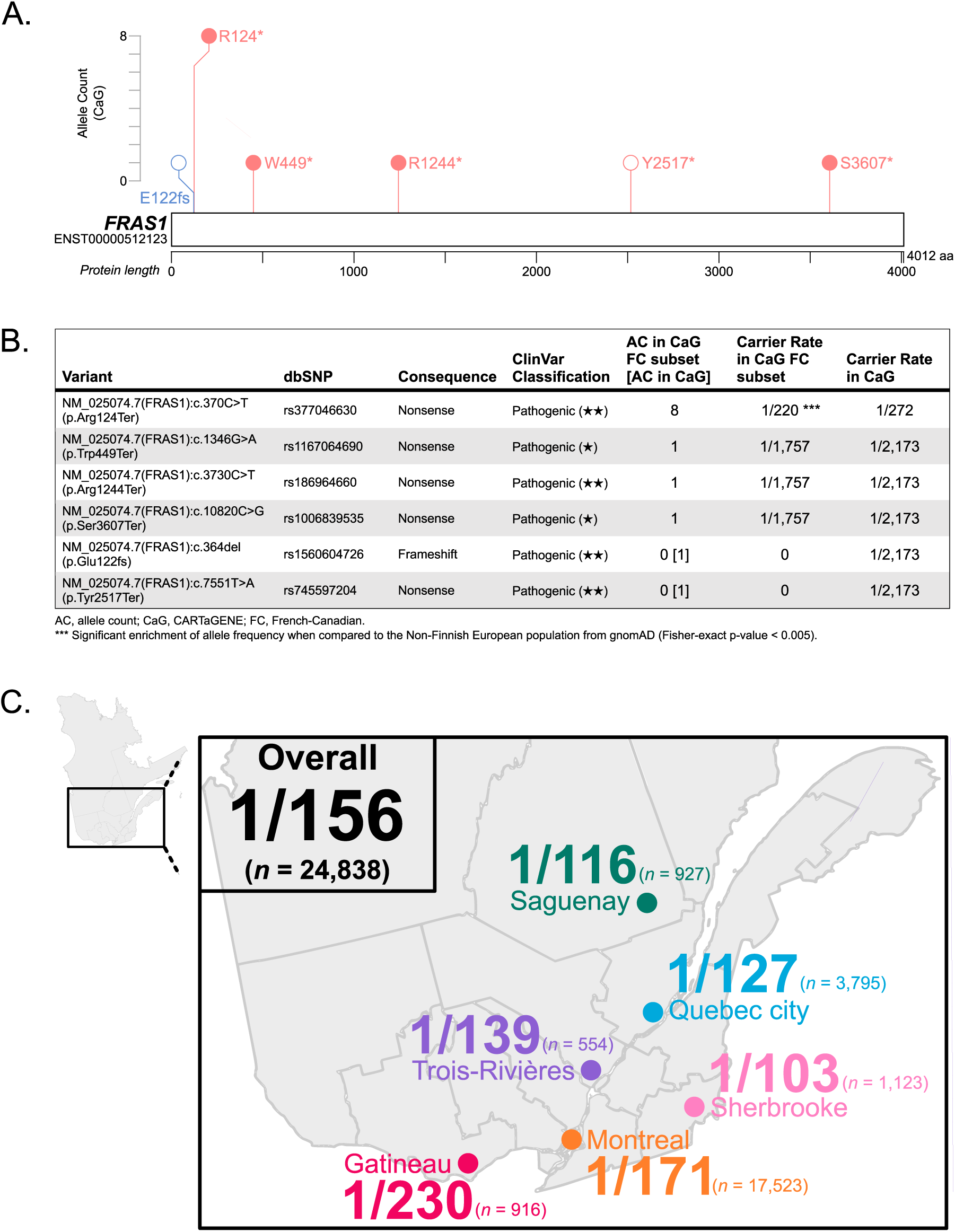
Fraser Syndrome Pathogenic Variants Identified in the Population-based Cohort CARTaGENE (CaG). **A. Distribution of Pathogenic/Likely Pathogenic (P/LP) Variants in FRAS1 Identified in CaG**. Genome Sequencing data included 2,173 individuals, of whom 1,756 were of French-Canadian (FC) genetic ancestry. Variants were annotated using the Variant Effect Predictor (VEP) tool (version 112) on the GRCh38 reference genome and mapped to canonical transcripts. Nonsense variants are depicted in red, and frameshift variants in blue. Variants identified in individuals of FC genetic ancestry are indicated with filled circles. The image was modified from Protein Paint (https://proteinpaint.stjude.org).^27^ **B. Summary of P/LP Variants Identified in CaG.** Genome sequencing data was used to identify P/LP variants. Allele counts (AC) and carrier rates are reported for the FC subset and the total CaG cohort. Carrier rates were calculated as the proportion of individuals carrying the variant. Bias-corrected odds ratio and Fisher-exact test were calculated from allele frequencies in the CaG FC subset and in the Non-Finnish European population from gnomAD (v.2.1.1). Additional information can be found in the Supplementary material (*S3. CARTaGENE Genome Sequencing Variants*). **C.** Regional Carrier Frequency of the FRAS1 p.(Arg124Ter) Variant in Quebec. Carriers were identified among 24,838 unrelated individuals of European genetic ancestry with genotyping data linked to specific metropolitan areas (Montreal, Quebec city, Sherbrooke, Saguenay, Trois-Rivières, Gatineau) where CaG participants were recruited. *n*, number of individuals sampled in each region.

To test if the frequency of pathogenic variants was higher in the French-Canadian population, we compared AF with those reported in the NFE population from gnomAD. We confirmed that one variant, *FRAS1* p.(Arg124Ter), is significantly enriched in both cohorts: in the CaG population (WGS dataset) with frequency approximately 21-fold higher than NFE (bias-corrected OR = 21.05; p-value = 1.12E-07), and in the Quebec IBD cohort at approximately 17-fold (bias-corrected OR = 17.38; p-value = 2.06E-06).

### Regional carrier frequency

AF geographic distribution was inferred using genotyping data from 24,838 unrelated European ancestry CaG participants sampled across six metropolitan areas: Montreal (*n* = 17,523), Quebec city (*n* = 3,795), Sherbrooke (*n* = 1,123), Saguenay (*n* = 927), Gatineau (*n* = 916), and Trois-Rivières (*n* = 554). The carrier rates presented here are minimal estimates, since only the *FRAS1* p.(Arg124Ter) variant was present in CaG genotyping array data. Overall, we found 1 in 156 individuals were carriers, with the highest regional concentrations observed in Sherbrooke (1/103), followed by Saguenay (1/117) and Quebec city (1/127) (Figure 1c).

### Estimated birth prevalence

We estimated the birth prevalence of FS from genomic data and reported cases, independently. Assuming Hardy-Weinberg equilibrium, we used the carrier frequency of disease-causing variants (*q*) to calculate the expected frequency of FS cases (*q*^2^). In the Quebec IBD cohort, based on the carrier frequency of *FRAS1* p.(Arg124Ter) variant in 2,323 individuals, the minimal estimated birth prevalence would be 0.23 per 100,000 births (95% CI [0.045, 0.87]) or 1 in 440,138. In CaG French-Canadian subset, we expect a birth prevalence of 0.98 per 100,000 (95% CI [0.28, 2.93]) or 1 in 101,935 births. In all of CaG, representing most of the Quebec population, the estimated birth prevalence is 0.89 per 100,000 (95% CI [0.28, 2.46]) or 1 in 111,855 births.

At CHU Ste-Justine, a tertiary care center covering 40% of the pediatric population in Quebec, four cases of self-reported French-Canadian ancestry were identified during the 10 years under review. This corresponds to a minimal estimated birth prevalence of 1.34 per 100,000 or 1 in 74,662 births. Including all cases (*n* = 5) and births (*n* = 373,310) irrespective of ancestry, we observed the same birth prevalence.

### Clinical characteristics

Table 1 presents clinical and genetic characteristics of the five FS patients identified in five unrelated families. Four were of self-reported French-Canadian ancestry and one of African ancestry. Among these cases, four resulted in termination of pregnancy due to fetal anomaly (TOPFA), and one died shortly after birth. Diagnoses were confirmed postnatally through clinical examination and genetic testing. Among all cases, five causal variants were identified: three nonsense, one missense, and one splice site variant, detected in homozygous (3/5) or compound heterozygous (2/5) state. The *FRAS1* p.(Arg124Ter) variant was the most common, accounting for 60% of disease-causing alleles (6/10), and the only one found in homozygous state.

**Table 1.** Clinical characteristics of five Fraser syndrome cases identified in one mother-child tertiary care center in Quebec over a decade.

|  | Case no. 1 | Case no. 2 | Case no. 3 | Case no. 4 | Case no. 5 |
| --- | --- | --- | --- | --- | --- |
| *Demographics* |  |  |  |  |  |
| Variants | *FRAS1*<br>p.Arg124Ter /<br>*FRAS1*<br>p.Arg124Ter | *FRAS1*<br>p.Arg124Ter /<br>*FRAS1*<br>p.Arg124Ter | *FRAS1*<br>p.Arg124Ter /<br>*FRAS1*<br>p.Arg124Ter | *FRAS1*<br>p.Trp449Ter /<br>*FRAS1*<br>c.2722+1G>A | *FRAS1*<br>p.Ile1951Thr /<br>*FRAS1*<br>p.Arg1709Ter |
| Sex | XX | XY | XX | XY | XX |
| Age at death | TOPFA | TOPFA | TOPFA | TOPFA | Neonatal |
| Origin of parents <sup>†</sup> | French-Canadian | French-Canadian | French-Canadian | French-Canadian | African ancestry |
| *Clinical features* |  |  |  |  |  |
| **Cryptophthalmos** | + | + | + | - | + |
| **Syndactyly** | - | + | + | + | + |
| **Urinary tract abnormalities** |  |  |  |  |  |
| Renal agenesis, or<br>Renal hypoplasia, or<br>Megacystis | + | + | + | + | + |
| **Ambiguous genitalia** |  |  |  |  |  |
| Ambiguous external<br>genitalia, or<br>Clitoromegaly, or<br>Vaginal atresia | + | - | + | - | + |
| **Laryngeal and tracheal**<br>**anomalies** | + | + | + | + | + |
| **Anorectal defects** | + | - | - | + | - |
| **Dysplastic ears** | + | + | + | + | + |
| **Skull ossification defects** | - | + | - | + | + |
| **Umbilical abnormalities** |  |  |  |  |  |
| Umbilical hernia, or<br>Omphalocele, or<br>Low-set umbilicus | + | - | + | - | - |
| **Nasal anomalies** | + | + | + | + | + |

Clinical features are presented according to the modified diagnostic criteria emitted by van Haelst et al. (2007). These include six major manifestations (syndactyly, cryptophthalmos spectrum, urinary tract abnormalities, ambiguous genitalia, laryngeal and tracheal anomalies, and positive family history) and five minor features (anorectal defects, dysplastic ears, skull ossification defects, umbilical abnormalities, and nasal anomalies) [van Haelst 2007]. A diagnosis of FS can be made if a patient presents either three major, or two major and two minor, or one major and three minor diagnostic criteria [van Haelst 2007]. In our study, all cases met at least three major and three minor criteria. Additional information on variants can be found in the supplementary materials (S5. Patient Variants).
**Bold:** major criteria; **bold and italics:** minor criteria.
TOPFA, termination of pregnancy due to fetal anomaly; w, weeks.
<sup>†</sup> Self-declared

Clinical manifestations are presented according to the modified diagnostic criteria emitted by van Haelst et al. (Table 1).^5^ Cryptophthalmos and syndactyly were each found in 80% of cases (4/5). All patients presented with urinary tract abnormalities, laryngeal and tracheal anomalies, dysplastic ears, and nasal anomalies. We highlight phenotypic heterogeneity, even among cases with the same genotype (patients 1, 2, and 3).

## DISCUSSION

FS is an autosomal recessive congenital malformation disorder with distinctive features. Here we show the unique genetic architecture of this syndrome in Quebec, with the *FRAS1* p.(Arg124Ter) pathogenic variant accounting for a majority of pathogenic alleles in French-Canadian FS probands (75%) as well as in healthy individuals of French-Canadian genetic ancestry from two independent cohorts: the Quebec IBD cohort (100%) and the population-based cohort CaG (73%). Based on these findings, we propose that *FRAS1* p.(Arg124Ter) is a candidate French-Canadian pathogenic founder variant.

In this study, the same homozygous *FRAS1* variant p.(Arg124Ter) was identified independently in three probands from unrelated French-Canadian families. In the literature, it is reported in only two other FS-affected individuals: a homozygous fetus (TOPFA) born to first-cousin parents from Turkey,^20^ and an 8-year-old heterozygous individual (with no disease-causing variant identified on the second allele) of unknown ethnicity.^21^ When we compare these reports with patients from our case series, we observe that the homozygous genotype p.(Arg124Ter) caused severe malformations and all parents opted to terminate the pregnancy.

Out of the four other causal variants identified in our case series, only *FRAS1* p.(Trp449Ter) was also present in CaG genome sequencing data. In a compound heterozygous state with the *FRAS1* c.2722+1G>A splice donor variant, it resulted in TOPFA at 16 weeks of gestation (Patient 4). Although many pathogenic variants in the *FRAS1*, *FREM2*, and *GRIP1* genes have been reported in the literature, most are private or family-specific. Only one variant has been described as possibly linked to a founder effect: *FRAS1* c.6963_6964dup, identified in two unrelated Polish families with FS who were homozygous for this pathogenic variant.^22^

Major anomalies are usually detected prenatally via ultrasound during the second trimester.^23^ Specific antenatal findings that suggest a clinical diagnosis of FS include cryptophthalmos and congenital high airway obstruction sequence (CHAOS). Oligohydramnios hampers the ascertainment of some manifestations, but its presence in combination with renal agenesis and CHAOS should still prompt the consideration of this diagnosis.^23^ In most cases, however, FS is diagnosed through clinical examination and/or perinatal autopsy when a patient presents either three major, or two major and two minor, or one major and three minor diagnostic criteria.^5,23^ In our study, all cases met at least three major and three minor criteria. Syndactyly and cryptophthalmos, considered to be the hallmark features of this syndrome, were each present in 80% (4/5) of cases. A better understanding of the antenatal presentation and phenotypic variability could potentially help healthcare providers offer genetic counselling. This is especially relevant in founder populations, where the presence of an enriched variant can affect the approach to molecular genetic testing.

The French-Canadians represent a founder population genetically structured at the regional scale (Bhérer et al., medRxiv 2025; preprint PMID: 40791678).^24^ So far, more than 30 disorders are known to bear a founder effect signature.^25,26^ Our results demonstrate enrichment in frequency of an otherwise very rare FS disease-causing variant, aligning with the founder effect. This underscores the importance of understanding pathogenic variants at elevated frequencies in founder populations to improve clinical practice and healthcare policies.

In our previous work involving exome sequence data on 2,323 unrelated French-Canadian individuals from the Quebec IBD cohort, the *FRAS1* p.(Arg124Ter) variant was found to be significantly enriched, with frequency approximately 17-fold higher than NFE (bias-corrected OR = 17.38; p-value = 2.06E-06). We replicated these results in the CaG cohort, where we observed a 21-fold enrichment of the same variant (bias-corrected OR = 21.05; p-value = 1.12E-07). From the carrier frequency of pathogenic variants in the Quebec IBD cohort and CaG, we estimated the minimal birth prevalence of FS in French Canadians to be 0.23 per 100,000 births (95% CI [0.045, 0.87]) and 0.98 per 100,000 births (95% CI [0.28, 2.93]), respectively. Independently, the birth prevalence observed from reported cases over a 10-year review period at CHU Sainte-Justine was found to be 1.34 per 100,000 births. We note that these rates are higher than the 0.20 per 100,000 births (1 in 495,633) reported in Europe by Barisic et al., with the highest regional mean prevalence being 0.43 per 100,000 births (1 in 230,695) in Western Europe.^7^ European cases had a high rate of consanguinity (27%), all from the Western region, which could explain the slight overlap with the lower bound of the 95% CI of our estimates.^7^

Moreover, using data on more than 20,000 samples from across Quebec, we observed a higher carrier frequency in some regions, with the highest estimate seen in Sherbrooke (1/103). This is in accordance with the reported fine-scale regional population structure of the French Canadians, with certain Mendelian diseases showing regional concentrations.^24^ Only a few other Mendelian diseases have been identified as potential founder effects in southeastern

Quebec, and few prior studies have reported regional carrier rates across Quebec. Most have focused on the Saguenay–Lac-Saint-Jean region or provided estimates using patient-based cohorts. In contrast, our analyses leverage two datasets with large sample sizes and province-wide recruitment strategy that allow for unprecedented power to detect regional variation in AF. Although the French-Canadian founder population is well-described in the literature, our results demonstrate that there are still novel prevalent Mendelian diseases to discover in the population.

Our study has some limitations. We can only estimate minimum birth prevalence since some true pathogenic variants are unknown, and therefore not considered. Our samples could also be under-powered to detect extremely rare variants, which is plausible given that 3/5 variants identified in FS cases were not found in either Quebec IBD cohort or CaG genome sequencing data. Regional carrier frequencies are also likely under-estimated since some pathogenic variants in genome sequencing data were absent in genotyping array data. The Quebec IBD cohort is comprised of over 2,000 French-Canadian individuals from the province of Quebec, but may not represent the exact regional distribution of the current population (Bhérer et al., medRxiv 2025; preprint PMID: 40791678). Recruitment for the CaG cohort was done in metropolitan areas, and therefore does not reflect the entire Quebec population (McClelland et al., medRxiv 2025; preprint PMID: 40463523).^10^ Lastly, the reliance on a single tertiary care center’s data may introduce an upward bias in our birth prevalence estimate, whereas a larger proportion of at-risk pregnancies are followed at CHU Sainte-Justine Hospital. Nonetheless, these caveats should have minimal impact on our conclusions since our findings from three different approaches provide consistent evidence of an elevated frequency of the *FRAS1* p.(Arg124Ter) variant. Indeed, we find multiple carriers of this pathogenic variant in each cohort (7/4,644 alleles in the Quebec IBD cohort and 8/3,512 alleles in CaG genomes) and ¾ homozygous individuals of French-Canadian ancestry diagnosed with FS, which is in line with a candidate founder variant.

In conclusion, our study provides valuable insights into the prevalence of FS in the French-Canadian population. The identification of *FRAS1* p.(Arg124Ter) as a candidate pathogenic founder variant highlights the value of population-based cohorts for studying the genetic epidemiology of rare diseases, which could help plan genetic services.

## DATA AVAILABILITY

The CaG individual-level genomic data is available from https://cartagene.qc.ca/en/researchers/access-request.html after scientific and ethical review. The variant-level summary statistics for pathogenic variants in the Quebec IBD cohort are published and publicly available (https://www.medrxiv.org/node/1028169.external-links.html). Patient data is available upon request, pending relevant scientific and ethical review.

## ACKNOWLEDGEMENTS

We thank the patients and their families for their participation. We are also grateful to the study participants from the Quebec IBD cohort and CARTaGENE.

## FUNDING STATEMENT

C.B. holds a Junior 1 award from the Fonds de Recherche du Québec-Santé (FRQS). J.D.R. holds a Tier 1 Canada Research Chair (#230625) and is supported by US National Institutes of Health grant no. DK062432.

## AUTHOR CONTRIBUTIONS

Conceptualization: C.B., G.G., P.M.C., J.D.R; Data curation: G.G., J.P., D.T., M.S.D, V.C., C.B.; Formal analysis: G.G., J.P.; Funding acquisition: C.B., J.D.R., P.M.C; Investigation: G.G., P.M.C., M-A.D., A.K., J.P.; Methodology: G.G., C.B., D.A-B.; Project administration: C.B., P.M.C.; Resources: C.B., P.M.C., J.D.R, D.T.; Software: J.P., D.T., M.S.D, V.C.; Supervision: C.B., P.M.C., J.D.R; Visualization: G.G., J.P.; Writing-original draft: G.G., C.B., P.M.C.; Writing-review & editing: all authors.

## ETHICS DECLARATION

The retrospective chart review was performed in accordance with the CHU Sainte-Justine IRB-approved protocol number 4072. Patient data was de-identified. Analysis of CaG data was approved by the ethical review board from the Faculty of Medicine and Health Sciences at McGill (IRB Number A07-M45-21B). Study protocols for the Quebec IBD cohort were approved by MHI Institutional Ethics Committee “Comité d’éthique de la recherche et du développement des nouvelles technologies” (IBD Genetics 2005-23 (05-813) and MTT : MP- 02-2017-7170, 2017-2202) and the Research Ethics Office of the Faculty of Medicine and Health Sciences at McGill University, Canada (IRB Number A01-M04-21A).

## CONFLICT OF INTEREST

Disclosure: C.B. serves as advisor for Medeloop Inc. and holds shares in the company. The remaining authors declare no competing interests.

## SUPPLEMENTARY MATERIAL

**Supplementary** Table 1 **(S1). Quebec IBD Exome Sequencing Variants.** 25 variants in three genes (*FRAS1*, *FREM2* and *GRIP1*) were classified as pathogenic, likely pathogenic, or had conflicting interpretations of pathogenicity according to ClinVar. Only the *FRAS1* c.370C>T (p.Arg124Ter) was interpreted as pathogenic after manual curation.

**Supplementary** Table 2 **(S2). Quebec IBD VariantValidator.** Variants identified in the Quebec IBD cohort were validated using *VariantValidator*, a variant description validation software.^28^

**Supplementary** Table 3 **(S3). CARTaGENE Genome Sequencing Variants.** 43 variants in three genes (*FRAS1*, *FREM2* and *GRIP1*) were classified as pathogenic, likely pathogenic, or had conflicting interpretations of pathogenicity according to ClinVar.

**Supplementary** Table 4 **(S4). CARTaGENE VariantValidator.** Variants identified in the CARTaGENE were validated using *VariantValidator*, a variant description validation software.^28^

**Supplementary** Table 5 **(S5). Proband Variants.** Variants ascertained in five probands diagnosed with Fraser Syndrome between 2013 and 2023 at CHU Sainte-Justine in Quebec.

**Supplementary** Table 6 **(S6). Proband VariantValidator.** Variants identified in probands diagnosed with Fraser Syndrome between 2013 and 2023 at CHU Sainte-Justine in Quebec were validated using *VariantValidator*, a variant description validation software.^2^

## Notes

### Author Declarations

The retrospective chart review was performed in accordance with the CHU Sainte-Justine IRB-approved protocol number 4072. Patient data was de-identified. Analysis of CARTaGENE data was approved by the ethical review board from the Faculty of Medicine and Health Sciences at McGill (IRB Number A07-M45-21B). Study protocols for the Quebec IBD cohort were approved by MHI Institutional Ethics Committee Comité d'éthique de la recherche et du développement des nouvelles technologies (IBD Genetics 2005-23 (05-813) and MTT : MP-02-2017-7170, 2017-2202) and the Research Ethics Office of the Faculty of Medicine and Health Sciences at McGill University, Canada (IRB Number A01-M04-21A).

## REFERENCES

1. McGregor, L., Makela, V., Darling, S. M., Vrontou, S., Chalepakis, G., Roberts, C., Smart, N., Rutland, P., Prescott, N., Hopkins, J., Bentley, E., Shaw, A., Roberts, E., Mueller, R., Jadeja, S., Philip, N., Nelson, J., Francannet, C., Perez-Aytes, A., Megarbane, A., … Scambler, P. J. (2003). Fraser syndrome and mouse blebbed phenotype caused by mutations in FRAS1/Fras1 encoding a putative extracellular matrix protein. Nature genetics, 34(2), 203–208. 10.1038/ng1142

2. Jadeja, S., Smyth, I., Pitera, J. E., Taylor, M. S., van Haelst, M., Bentley, E., McGregor, L., Hopkins, J., Chalepakis, G., Philip, N., Perez Aytes, A., Watt, F. M., Darling, S. M., Jackson, I., Woolf, A. S., & Scambler, P. J. (2005). Identification of a new gene mutated in Fraser syndrome and mouse myelencephalic blebs. Nature genetics, 37(5), 520–525. 10.1038/ng1549

3. Vogel, M. J., van Zon, P., Brueton, L., Gijzen, M., van Tuil, M. C., Cox, P., Schanze, D., Kariminejad, A., Ghaderi-Sohi, S., Blair, E., Zenker, M., Scambler, P. J., Ploos van Amstel, H. K., & van Haelst, M. M. (2012). Mutations in GRIP1 cause Fraser syndrome. Journal of medical genetics, 49(5), 303–306. 10.1136/jmedgenet-2011-100590

4. Kiyozumi, D., Sugimoto, N., & Sekiguchi, K. (2006). Breakdown of the reciprocal stabilization of QBRICK/Frem1, Fras1, and Frem2 at the basement membrane provokes Fraser syndrome-like defects. Proceedings of the National Academy of Sciences of the United States of America, 103(32), 11981–11986. 10.1073/pnas.0601011103

5. van Haelst, M. M., Scambler, P. J., Fraser Syndrome Collaboration Group, & Hennekam, R. C. (2007). Fraser syndrome: a clinical study of 59 cases and evaluation of diagnostic criteria. American journal of medical genetics. Part A, 143A(24), 3194– 3203. 10.1002/ajmg.a.31951

6. Slavotinek, A. M., & Tifft, C. J. (2002). Fraser syndrome and cryptophthalmos: review of the diagnostic criteria and evidence for phenotypic modules in complex malformation syndromes. Journal of medical genetics, 39(9), 623–633. 10.1136/jmg.39.9.623

7. Barisic, I., Odak, L., Loane, M., Garne, E., Wellesley, D., Calzolari, E., Dolk, H., Addor, M. C., Arriola, L., Bergman, J., Bianca, S., Boyd, P. A., Draper, E. S., Gatt, M., Haeusler, M., Khoshnood, B., Latos-Bielenska, A., McDonnell, B., Pierini, A., Rankin, J., … Tenconi, R. (2013). Fraser syndrome: epidemiological study in a European population. American journal of medical genetics. Part A, *161A*(5), 1012– 1018. 10.1002/ajmg.a.35839

8. EUROCAT. (2018). *EUROCAT Guide 1.4: Instruction for the registration of congenital anomalies*. https://eu-rd-platform.jrc.ec.europa.eu/sites/default/files/Full_Guide_1_4_version_28_DEC2018.pdf

9. Gregg, A. R., Aarabi, M., Klugman, S., Leach, N. T., Bashford, M. T., Goldwaser, T., Chen, E., Sparks, T. N., Reddi, H. V., Rajkovic, A., Dungan, J. S., & ACMG Professional Practice and Guidelines Committee (2021). Screening for autosomal recessive and X-linked conditions during pregnancy and preconception: a practice resource of the American College of Medical Genetics and Genomics (ACMG). Genetics in medicine : official journal of the American College of Medical Genetics, 23(10), 1793–1806. 10.1038/s41436-021-01203-z

10. Awadalla, P., Boileau, C., Payette, Y., Idaghdour, Y., Goulet, J. P., Knoppers, B., Hamet, P., Laberge, C., & CARTaGENE Project (2013). Cohort profile of the CARTaGENE study: Quebec’s population-based biobank for public health and personalized genomics. International journal of epidemiology, 42(5), 1285–1299. 10.1093/ije/dys160

11. *CARTaGENE*. (n.d.). https://cartagene.qc.ca/index.html

12. Mclaren, W., Gil, L., Hunt, S. E., Riat, H. S., Ritchie, G. R. S., Thormann, A., Flicek, P., & Cunningham, F.. (2016). The Ensembl Variant Effect Predictor. Genome Biology, 17(1). 10.1186/s13059-016-0974-4

13. Karczewski, K. J., Francioli, L. C., Tiao, G., Cummings, B. B., Alföldi, J., Wang, Q., Collins, R. L., Laricchia, K. M., Ganna, A., Birnbaum, D. P., Gauthier, L. D., Brand, H., Solomonson, M., Watts, N. A., Rhodes, D., Singer-Berk, M., England, E. M., Seaby, E. G., Kosmicki, J. A., Walters, R. K., … MacArthur, D. G. (2020). The mutational constraint spectrum quantified from variation in 141,456

14. humans. *Nature*, 581(7809), 434–443. 10.1038/s41586-020-2308-7

15. Boissel, S., Fallet-Bianco, C., Chitayat, D., Kremer, V., Nassif, C., Rypens, F., Delrue, M. A., Dal Soglio, D., Oligny, L. L., Patey, N., Flori, E., Cloutier, M., Dyment, D., Campeau, P., Karalis, A., Nizard, S., Fraser, W. D., Audibert, F., Lemyre, E., Rouleau, G. A., … Michaud, J. L. (2018). Genomic study of severe fetal anomalies and discovery of GREB1L mutations in renal agenesis. Genetics in medicine : official journal of the American College of Medical Genetics, 20(7), 745– 753. 10.1038/gim.2017.173

16. Institut de la statistique du Québec. (2024, July 25). *Births and Birth Rate, Québec, 1900-2023 (in French only)*. Institut De La Statistique Du Québec. https://statistique.quebec.ca/en/document/births-quebec/tableau/births-and-birth-rate-quebec

17. Institut de la statistique du Québec. (2024a, May 8). *Stillbirths and Infant Deaths by Duration, Québec, 1976-2023 (in French only)*. Institut De La Statistique Du Québec. https://statistique.quebec.ca/en/document/stillbirths-and-infant-deaths/tableau/stillbirths-and-infant-deaths-by-duration-quebec

18. Statistics Canada. 2011 Census of Population http://www.statcan.gc.ca, 2012.

19. Université de Montréal - Faculté de Médecine. (2024, June 7). *Université de Montréal - Faculté de Médecine*. https://medecine.umontreal.ca/faculte/reseau-en-sante/#~:text=Tous%20les%20h%C3%B4pitaux%20et%20centres,de%20la%20population%20du%20Qu%C3%A9bec

20. Rivas, M. A., Avila, B. E., Koskela, J., Huang, H., Stevens, C., Pirinen, M., Haritunians, T., Neale, B. M., Kurki, M., Ganna, A., Graham, D., Glaser, B., Peter, I., Atzmon, G., Barzilai, N., Levine, A. P., Schiff, E., Pontikos, N., Weisburd, B., … Daly, M. J.. (2018). Insights into the genetic epidemiology of Crohn’s and rare diseases in the Ashkenazi Jewish population. PLOS Genetics, 14(5), e1007329. 10.1371/journal.pgen.1007329

21. Ogur, G., Zenker, M., Tosun, M., Ekici, F., Schanze, D., Ozyilmaz, B., & Malatyalioglu, E. (2011). Clinical and molecular studies in two families with Fraser syndrome: a new FRAS1 gene mutation, prenatal ultrasound findings and implications for genetic counselling. *Genetic counseling (Geneva*, Switzerland), 22(3), 233–244.

22. Kunz, F., Kayserili, H., Midro, A., de Silva, D., Basnayake, S., Güven, Y., Borys, J., Schanze, D., Stellzig-Eisenhauer, A., Bloch-Zupan, A., & Zenker, M. (2020). Characteristic dental pattern with hypodontia and short roots in Fraser syndrome. American journal of medical genetics. Part A, 182(7), 1681–1689. 10.1002/ajmg.a.61610

23. Midro, A. T., Stasiewicz-Jarocka, B., Borys, J., Hubert, E., Skotnicka, B., Hassmann- Poznańska, E., Sierpińska, T., Panasiuk, B., Schanze, D., & Zenker, M. (2020). Two unrelated families with variable expression of Fraser syndrome due to the same pathogenic variant in the FRAS1 gene. American journal of medical genetics. Part A, 182(4), 773–779. 10.1002/ajmg.a.61495

24. Tessier, A., Sarreau, M., Pelluard, F., André, G., Blesson, S., Bucourt, M., Dechelotte, P., Faivre, L., Frébourg, T., Goldenberg, A., Goua, V., Jeanne-Pasquier, C., Guimiot, F., Laquerriere, A., Laurent, N., Lefebvre, M., Loget, P., Maréchaud, M., Mechler, C., Perez, M. J., … Guerrot, A. M. (2016). Fraser syndrome: features suggestive of prenatal diagnosis in a review of 38 cases. Prenatal diagnosis, 36(13), 1270–1275. 10.1002/pd.4971

25. Bherer, C., Labuda, D., Roy-Gagnon, M. H., Houde, L., Tremblay, M., & Vézina, H. (2011). Admixed ancestry and stratification of Quebec regional populations. American journal of physical anthropology, 144(3), 432–441. 10.1002/ajpa.21424

26. Bchetnia, M., Bouchard, L., Mathieu, J., Campeau, P. M., Morin, C., Brisson, D., Laberge, A. M., Vézina, H., Gaudet, D., & Laprise, C. (2021). Genetic burden linked to founder effects in Saguenay-Lac-Saint-Jean illustrates the importance of genetic screening test availability. Journal of medical genetics, 58(10), 653–665. 10.1136/jmedgenet-2021-107809

27. Laberge, A. M., Michaud, J., Richter, A., Lemyre, E., Lambert, M., Brais, B., & Mitchell, G. A. (2005). Population history and its impact on medical genetics in Quebec. Clinical genetics, 68(4), 287–301. 10.1111/j.1399-0004.2005.00497.x

28. Zhou, X., Edmonson, M. N., Wilkinson, M. R., Patel, A., Wu, G., Liu, Y., Li, Y., Zhang, Z., Rusch, M. C., Parker, M., Becksfort, J., Downing, J. R., & Zhang, J. (2015). Exploring genomic alteration in pediatric cancer using ProteinPaint. Nature Genetics, 48(1), 4-6. 10.1038/ng.3466

29. Freeman, PJ, Hart, RK, Gretton, LJ, Brookes, AJ, Dalgleish, R. VariantValidator: Accurate validation, mapping and formatting of sequence variation descriptions. Human Mutation. 2018;39:61–68. 10.1002/humu.23348

